# The tongue biofilm metatranscriptome identifies metabolic pathways associated with halitosis and its prevention

**DOI:** 10.1101/2021.11.09.21266067

**Authors:** M. Carda-Diéguez, B.T. Rosier, S. Lloret, C. Llena, A. Mira

## Abstract

Halitosis is an oral condition caused by an increase in the concentration of volatile sulfur compounds (VSCs), such as methyl mercaptan and hydrogen sulfide, generated as a consequence of bacterial metabolism on the tongue biofilm. Microbial communities on the tongue of halitosis patients have been studied by bacterial culture, 16S rRNA taxonomic studies and metagenomics. However, there are currently no reports on the microbial gene-expression profiles. In this study, we performed RNAseq of tongue coating samples from control individuals and halitosis patients with different levels and composition of VSCs, as determined by gas chromatography. In this metatranscriptomic study, the activity of *Streptococcus, Veillonella* and *Rothia* species was associated with halitosis-free individuals while *Prevotella, Fusobacterium* and *Leptotrichia* species were associated with halitosis. Although methyl mercaptan is considered an indicator of halitosis, the metatranscriptome of patients in which only this VSC was present in elevated levels was similar to that of halitosis-free individuals. *Veillonella dispar, Streptococcus parasanguinis* and *Rothia mucilaginosa* were over-represented in halitosis-free communities in comparison to the rest of the groups, suggesting that these species could be used as a halitosis-free biomarkers. In contrast, the abundance of *Prevotella shahi* and *Fusobacterium nucleatum* were significantly higher when hydrogen sulfide concentration was over the established halitosis-threshold, making these species putative halitosis biomarkers. Finally, gene expression profiles showed a significant over-expression of genes involved in L-cysteine and L-homocysteine synthesis in halitosis-free individuals and an over-expression of genes responsible for cysteine degradation into hydrogen sulfide in halitosis patients. In addition, nitrate reduction into nitrite was also over-expressed in halitosis-free patients. In conclusion, halitosis was associated with communities that degrade amino acids and reduce sulfide, whereas tongue communities that produce L-cysteine from hydrogen sulfide and that reduce nitrate were associated with the absence of halitosis. The latter could provide new strategies to treat this condition.

## Introduction

Halitosis is a highly prevalent condition characterized by oral malodor [1]. A Swedish study with more than 800 participants showed a 2% prevalence while in a Chinese population (2500 participants) the estimated proportion increased up to 27,5% [2,3]. A recent systematic revision estimated a 31.8% prevalence of halitosis after examining 548 publications [4]. Depending on the origin of the oral malodor, halitosis can be differentiated into intra-oral (90% of cases), extra-oral and transient halitosis [5]. Several factors have been shown to affect bad breath, including necrotic pulpal exposure, deep carious lesions, specific food items, oral infections, periodontal disease, faulty restorations, reduced salivary flow, smoking and poor oral hygiene [6–10]. The oral malodor derives from the increase in the levels of several volatile sulfur compounds (VSCs), such as dimethyl sulfide (“DMS”, [CH3]_2_S), hydrogen sulfide (“HS”, H_2_S) and methyl mercaptan (“MM”, CH_3_SH). The last two have been unequivocally related to intra-oral halitosis and appear to be responsible for up to 90% of oral VSCs [11] whereas dimethyl sulfide is considered to be implicated mainly in extra-oral blood borne halitosis [12]. Current clinical literature establishes specific threshold values for halitosis diagnosis for H_2_S (112 ppb), CH_3_SH (26 ppb) and [CH_3_]_2_S (8 ppb) [13–15]. Previous epidemiological reports suggest that other odoriferous molecules could also be responsible or contribute to bad breath, including volatile aromatic compounds and (poly)amines, short/medium-chain fatty acids or organic acids, alcohols, volatile aliphatic compounds, aldehydes and ketones [16]. Diverse microbiological and epidemiological studies have shown that VSCs are mainly produced by oral microbiota degradation of cysteine, cystine and methionine, as well as tryptophan, arginine and lysine (see Bollen et al. 2012 for a review)[17]. Most intra-oral halitosis is associated with microbial activity on the tongue and it has been reported that tongue coating in halitosis patients is thicker than that in healthy individuals [18,19]. Periodontal pockets can also be a source of halitosis and it is interesting that tongue coating in periodontitis patients is 4 times more abundant compared to healthy individuals [20], suggesting that both diseases could be interconnected. It has also been shown that hydrogen sulfide has cytotoxic and pro-inflammatory properties under certain conditions, potentially contributing to inflammation and tissue damage in periodontitis [21,22] and therefore halitosis could have an effect on human health beyond the undesirable consequences of oral bad odor.

Microbial communities associated with caries, periodontitis and halitosis are known to have different compositions and functions in comparison to healthy individuals, as a consequence of a microbial dysbiosis [23,24]. For example, species from *Neisseria* and *Rothia* are commonly found in higher concentration in healthy individuals in comparison to those with oral disease [25]. When the tongue microbiota of halitosis patients or from those with higher tongue coating is compared to that of halitosis-free individuals, VSCs producers such as *Prevotella, Fusobacterium* and *Leptotrichia* are detected in higher proportions [5,26,27].

The relationship of tongue- and saliva-associated microbiotas with halitosis has been studied recently using high-throughput sequencing approaches. Ren and colleagues studied in 2016 the tongue- and saliva-associated microbiotas in Chinese children with halitosis [5] using 16S rRNA sequencing and shotgun metagenomics (direct, whole DNA sequencing) to study the changes in microbiota composition and functions. Similarly, Ye et al in 2019 used 16S rRNA sequencing in Chinese adults [28]. Seerangaiyan *et al*. used metabolomics to detect putative molecules associated with tongue coating [29]. However, there is currently no evidence that tongue-associated microbes are over-expressing genes important for VSCs production in halitosis patients and if there are other functions or metabolic pathways activated that could favor the disease. Thus, the application of current RNAseq technologies to oral samples, by which the total metatranscriptomic pool from microbial communities is analyzed [30,31], would allow the detection and quantification of all metabolic pathways that could be activated and repressed in halitosis patients.

In the current study, we have sequenced the total RNA of the microbial communities associated with the tongue coating to provide the first metatranscriptomic profile of tongue-associated biofilms under halitosis-free and halitosis conditions. In addition, we have differentiated halitosis patients according to the concentration of different VSCs separately, with the purpose of relating microbial activity to specific metabolic pathways, thereby shedding light to the underlying mechanisms of VSC production and to potential therapeutic approaches.

## Methods

### Study population

This study was performed at the University of Valencia Dental Clinic, managed by the Lluís Alcanyís Foundation (Valencia, Spain), and conducted according to the ethical principles of the Declaration of Helsinki of 2008. The study protocol was approved by the FISABIO-DGSP Ethical Committee (B1O2015-68711-R) and all participants signed an informed consent prior to sample donation. Potentially suitable individuals who visited the dental clinic for revision or treatment were invited to a screening for participation. The inclusion criteria for participants were between 18 and 65 years of age, systemically healthy, and presence of at least 20 teeth. Individuals were excluded if they suffered from any systemic disease (including liver or renal diseases, bronchial disorders, uncontrolled diabetes, gastroesophageal disease and reflux, cancer, autoimmune diseases, and anemia). Other exclusion criteria included pregnant or nursing women, as well as antibiotic treatment or regular antiseptic use in the last month. Participants who fulfilled the selection criteria received the following dietary instructions for the day of their visit, including avoidance of food with garlic, onion or strong spices, no alcohol consumption 24h before the visit, no coffee consumption in the 6h before the visit, no food 2 hours before the visit, no tongue brushing on the day of the visit, and avoidance of products with mint or perfume.

### VSCs measurements and patient classification

The concentrations of three VSCs (H_2_S, MM and DMS) in the breath of 83 individuals were measured (in ppb) using an Oralchroma portable gas chromatograph (**Figure 1 and Sup Table 1**). For the VSCs measurement, patients were asked to breath in a sterile syringe that was immediately injected into the OralChroma and measurements were taken according to the manufacturer’s instructions. Based on the measurements of the two main intraoral halitosis gases, H_2_S and MM, individuals were classified into four groups (**Figure 1**): a control group (Ctr) of halitosis-free individuals with both gasses below the halitosis-threshold (<112 ppb for H_2_S and <26 ppb for CH_3_SH, [13–15]) and three halitosis groups, namely the MM group (<112 ppb H_2_S and >26 ppb CH_3_SH), the H_2_S group (>112 ppb H_2_S and <26 ppb CH_3_SH), and the MM-HS group (>112 ppb H_2_S and >26 ppb CH_3_SH). In each group, the 10 individuals that best fulfilled the criteria (i.e., lowest or highest ppb of one or both of the gasses) were selected for RNAseq analysis.

**Figure 1.**
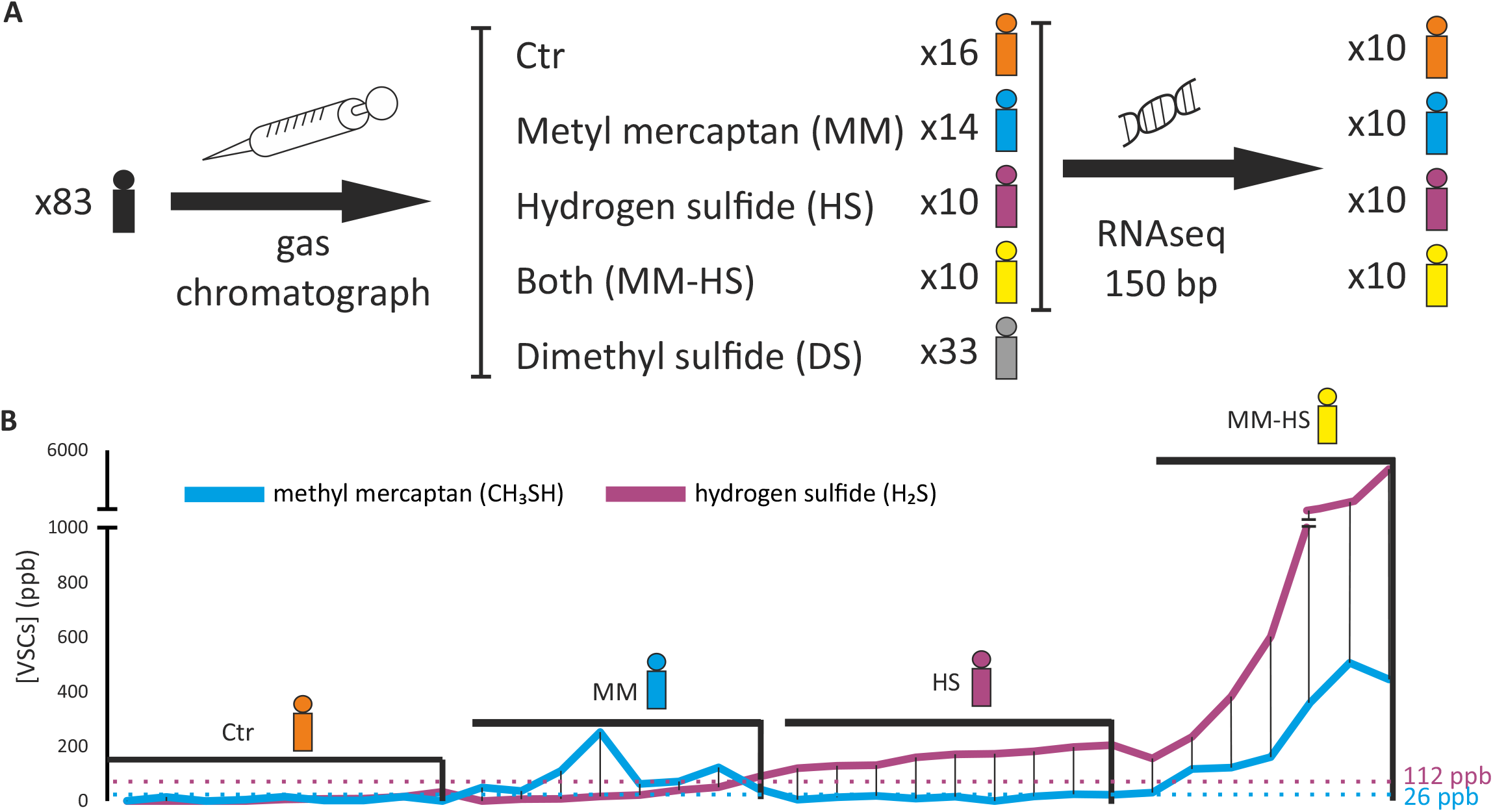
Study design and Volatile Sulfur Compounds (VSCs) concentrations in control (Ctr) and halitosis groups. **A**. Individuals were asked to breath into a syringe that was immediately introduced into an OralChroma gas chromatograph to measure VSCs concentrations (ppm). Individuals were allocated to five groups based on the levels of three VSCs (hydrogen sulfide - HS-, methyl mercaptan -MM-, and dimethyl sulfide -DMS-). Ten individuals per group were selected, corresponding to control individuals, and to the three intra-oral halitosis groups (MM, HS and MM+HS). Total RNA was extracted and sequenced by an Illumina Nextseq platform. The fifth group (DMS) contained 33 individuals that only had dimethyl sulfide above the established threshold (8 ppb), but this is generally considered an extra-oral gas and was not included in the metatranscriptome analysis. **B**, the concentrations of methyl mercaptan and hydrogen sulfide are shown for individuals in the Ctr, MM, HS and MM+HS groups. Note that only individuals for which the RNA was successfully sequenced are shown (N = 9 for Ctr, N = 8 for MM, N = 9 for HS, N = 7 for MM-HS). The established methyl mercaptan and hydrogen sulfide halitosis thresholds (112 and 26 ppb, respectively) are represented with dotted lines.

### Tongue microbiota sampling

Tongue coating samples (containing the tongue microbiota) were collected from the selected 40 participants from the middle and posterior sections of the tongue by scrapping with an autoclaved Heidemann spatula four times. The samples were put into 0.5 ml of RNAlater stabilizing solution (Invitrogen, Thermo Fisher Scientific) and directly frozen at -20°C until RNA extraction.

### Oral health assessment

A Periodontogram was used and a gingival index was determined following Löe et al. (1963) and Camelo-Castillo et al., (2015) [32,33]. The flow of unstimulated saliva and caries assessment following ICDAS classification system were registered following Ferrer et al. (2021) [34]. The color of the tongue coating (white, light yellow, dark yellow, brown), its extension (one, two or three thirds) and its width (0: absence of tongue biofilm; 1: thin layer, that allows visualization of papillae; 2: moderate layer, papillae are partially seen; 3: thick layer, papillae are not seen) were recorded. In addition, the individuals filled out a questionnaire with information about oral hygiene and dietary questions. For this study, the analyzed parameters included tongue brushing (yes/no and frequency), dental flossing (yes/no) and mouthwash usage (yes/no). Additionally, the analyzed dietary parameters included protein-rich diet (yes/no) and the consumption of garlic or onion (garlic/onion/both) and dairy products (yes/no).

### Sampling, RNA extraction and sequencing

The tongue coating samples were thawed on ice and nucleic acids were manually extracted using the Master Pure Complete DNA and RNA Purification Kit (Epicentre), following the manufacturer protocol, and resuspended in a final volume of 30 µl. Every step during RNA extraction was performed with RNase-free, low-retention Eppendorf tubes and RNase-free water (Sigma). The DNA+RNA extracted was treated 3 times with a Turbo DNA-free Kit (Ambion) at 37°C for 30 minutes each in order to eliminate DNA and keep RNA. DNA-free material was amplified (PCR) using universal 16S rRNA primers to confirm the absence of DNA traces [31]. Genomic DNA was used as positive control. The PCR program included 2 minutes at 94°C, 30 cycles (94°C 15’’, 55 °C 30’’ and 72°C 40’’) and a final amplification step 72 °C for 5 minutes. The absence of a DNA band was considered evidence of correct DNAse treatment. RNA concentration was measured with Qubit RNA BR Assay Kit (ref:Q10210). Libraries and cDNA sequencing were performed at the FISABIO sequencing platform (Valencia, Spain) using NextSeq Illumina Technology (single pairs, High-input 1×150 bp) following the manufacturer’s instructions.

### Bioinformatic analysis

Reads were trimmed by quality and length using the PRINSEQ program [35]. Human and microbial ribosomal sequences were identified and separated by aligning the sequence dataset to the human genome (Bioproject PRJNA31257) and SILVA database, respectively [36]. Remaining reads were considered to be mRNA reads and were used for annotation. We used a manually curated version of the Human Oral Microbial Database (HOMD) [37] to annotate the mRNA reads. The abundance of active bacteria was studied using the number of reads aligned to bacterial ORFs from the HOMD. To eliminate the potential bias caused by differences in gene length and dataset size, the number of reads annotated to a gene were normalized by its length (Kbp) and the size (in Megabp) of the dataset. Thus, the level of gene expression was indicated as number of reads per gene Kbp per Mbp (RPKM).

We assigned a KEGG model to every Open Reading Frame (ORF) by aligning the amino acid dataset to the KEGG database [38] with HMMER [39]. The abundance of a fragment was considered to be equal to the number of sequences aligned into this fragment using BLAST [40]. The calculations for obtaining the abundance matrices and the comparative analyses (see below) were performed using custom R scripts (http://www.R-project.org/). Principal component and canonical correspondence correlation analyses were performed in order to display differences between the groups. DESeq2 test was used to calculate the significance of differences between groups [41]. Analyses were performed with the significance level of p<0.05 (significant p-value) or p<0.1 (statistical trend).

## Results

### Sequencing results

Due to low RNA quality hindering library construction or to a low number of quality-filtered reads, the final number of samples per group was 9, 8, 9 and 7 for *Ctr, MM, HS* and *MM-HS* groups, respectively (**Figure 1A and B**). The clinical parameters in each group are described in table 1. After quality-filtering, an average of 3.7×10^3^ Gbp were obtained per sample. On average, 0.1% of reads belonged to the host and 93.5% of reads were lost after ribosomal RNA elimination. Once filtered, an average of 0.7 million mRNA reads per sample were annotated (53%) and used for the metatranscriptomic analysis of the tongue microbiota.

**Table 1:** summary of clinical parameters in the healthy control group and halitosis groups

| Parameters |  |  | Control*<sup>1</sup><br>(n = 16) | MM<br>(n = 14) | HS<br>(n = 10) | MM-HS<br>(n = 10) | DMS<br>(n = 33) | All<br>(n = 83) |
| --- | --- | --- | --- | --- | --- | --- | --- | --- |
| Gender | Male | N (%) | 5 (31.3%) | 3 (21.4%) | 3 (30.0%) | 5 (50.0%) | 14 (42.4%) | 30 (36.1%) |
|  | Female | N (%) | 11 (68.8%) | 11 (78.6%) | 7 (70.0%) | 5 (50.0%) | 19 (57.6%) | 53 (63.9%) |
| Age (years) | Average (SD) |  | 47.0 (13.9) | 47.0 (13.6) | 34.9 (13.2) | 38.7 (16.4) | 46.9 (12.7) | 44.5 (14.0) |
| H2S (ppb)*<sup>2</sup> | Average (SD) |  | 29.2 (32.6) | 28.8 (26.1) | 170.23<br>(36.4) | 1237.38<br>(1428.4) | 41.1 (31.3) | 196.4<br>(614.0) |
| MM (ppb) | Average (SD) |  | 7.1 (6.3) | 72.5 (58.2) | 14.38 (7.7) | 280.52<br>(250.0) | 5.5 (6.4) | 51.3<br>(123.7) |
| DMS (ppb) | Average (SD) |  | 4.7 (2.4) | 23.2 (18.2) | 67.01<br>(79.4) | 79.75<br>(77.8) | 47.6 (35.8) | 41.4 (50.0) |
| Tongue colour | White | N (%) | 8 (50.0%) | 9 (64.3%) | 7 (70.0%) | 7 (70.0%) | 18 (54.5%) | 49 (59.0%) |
|  | Light yellow | N (%) | 3 (18.8%) | 1 (7.1%) | 0 (0.0%) | 0 (0.0%) | 4 (12.1%) | 8 (9.6%) |
|  | Dark yellow | N (%) | 5 (31.3%) | 4 (28.6%) | 3 (30.0%) | 3 (30.0%) | 11 (33.3%) | 26 (31.3%) |
| Tongue coating thickness | None | N (%) | 0 (0.0%) | 0 (0.0%) | 0 (0.0%) | 0 (0.0%) | 2 (6.1%) | 2 (2.4%) |
|  | Thin | N (%) | 9 (56.3%) | 7 (50.0%) | 8 (80.0%) | 3 (30.0%) | 14 (42.4%) | 41 (49.4%) |
|  | Moderate | N (%) | 5 (31.3%) | 7 (50.0%) | 1 (10.0%) | 7 (70.0%) | 16 (48.5%) | 36 (43.4%) |
|  | Thick | N (%) | 2 (12.5%) | 0 (0.0%) | 1 (10.0%) | 0 (0.0%) | 1 (3.0%) | 4 (4.8%) |
| Tongue coating extension | None | N (%) | 0 (0.0%) | 0 (0.0%) | 0 (0.0%) | 0 (0.0%) | 1 (3.0%) | 1 (1.2%) |
|  | <1/3 of tongue | N (%) | 8 (50.0%) | 6 (42.9%) | 5 (50.0%) | 2 (20.0%) | 8 (24.2%) | 29 (34.9%) |
|  | 1/3-2/3 of tongue | N (%) | 7 (43.8%) | 5 (35.7%) | 4 (40.0%) | 6 (60.0%) | 20 (60.6%) | 42 (50.6%) |
|  | >2/3 of tongue | N (%) | 1 (6.3%) | 3 (21.4%) | 1 (10.0%) | 2 (20.0%) | 4 (12.1%) | 11 (13.3%) |
| Frank cavitation*<sup>3</sup> | Yes | N (%) | 2 (12.5%) | 2 (14.3%) | 3 (30.0%) | 1 (10.0%) | 3 (9.1%) | 11 (13.3%) |
|  | No | N (%) | 14 (87.5%) | 12 (85.7%) | 7 (70.0%) | 9 (90.0%) | 30 (90.9%) | 72 (86.7%) |
| Bleeding any site | Yes | N (%) | 2 (12.5%) | 3 (21.4%) | 2 (20.0%) | 3 (30.0%) | 3 (9.1%) | 13 (15.7%) |
|  | No | N (%) | 14 (87.5%) | 11 (78.6%) | 8 (80.0%) | 7 (70.0%) | 30 (90.9%) | 70 (84.3%) |
| Periodontal pockets ≥4 mm any site | Yes | N (%) | 4 (25.0%) | 5 (35.7%) | 2 (20.0%) | 4 (40.0%) | 8 (24.2%) | 23 (27.7%) |
|  | No | N (%) | 12 (75.0%) | 9 (64.3%) | 8 (80.0%) | 6 (60.0%) | 25 (75.8%) | 60 (72.3%) |

\*<sup>1</sup>In the first row, the groups are shown (Control is the halitosis-free group, MM is the methyl mercaptan group, HS is the hydrogen sulfide group, MM-HS is the group with both previous gasses above the halitosis threshold, and DMS is the dimethyl sulfide group).
\*<sup>2</sup> H<sub>2</sub>S is hydrogen sulfide in parts per billion (ppb)
\*<sup>3</sup> Frank cavitation was considered any caries with ICDAS score $\geq 5$ and lost or fractured restoration

### Changes in active microbial proportions

According to the Canonical Correspondence Analysis (CCA) a significant clustering between groups was obtained at the species taxonomic composition (p=0.001) although a non-significant effect was detected by an ADONIS test (p=0.19), probably due to a high intragroup variation within the *Ctr* and *MM*-*HS* groups (**Figure 2A**). Inter-group differences were most apparent between the *Ctr* and the *HS* groups and the *Ctr* and the *MM-HS* groups, which were both separated by the CCA1 component.

**Figure 2.**
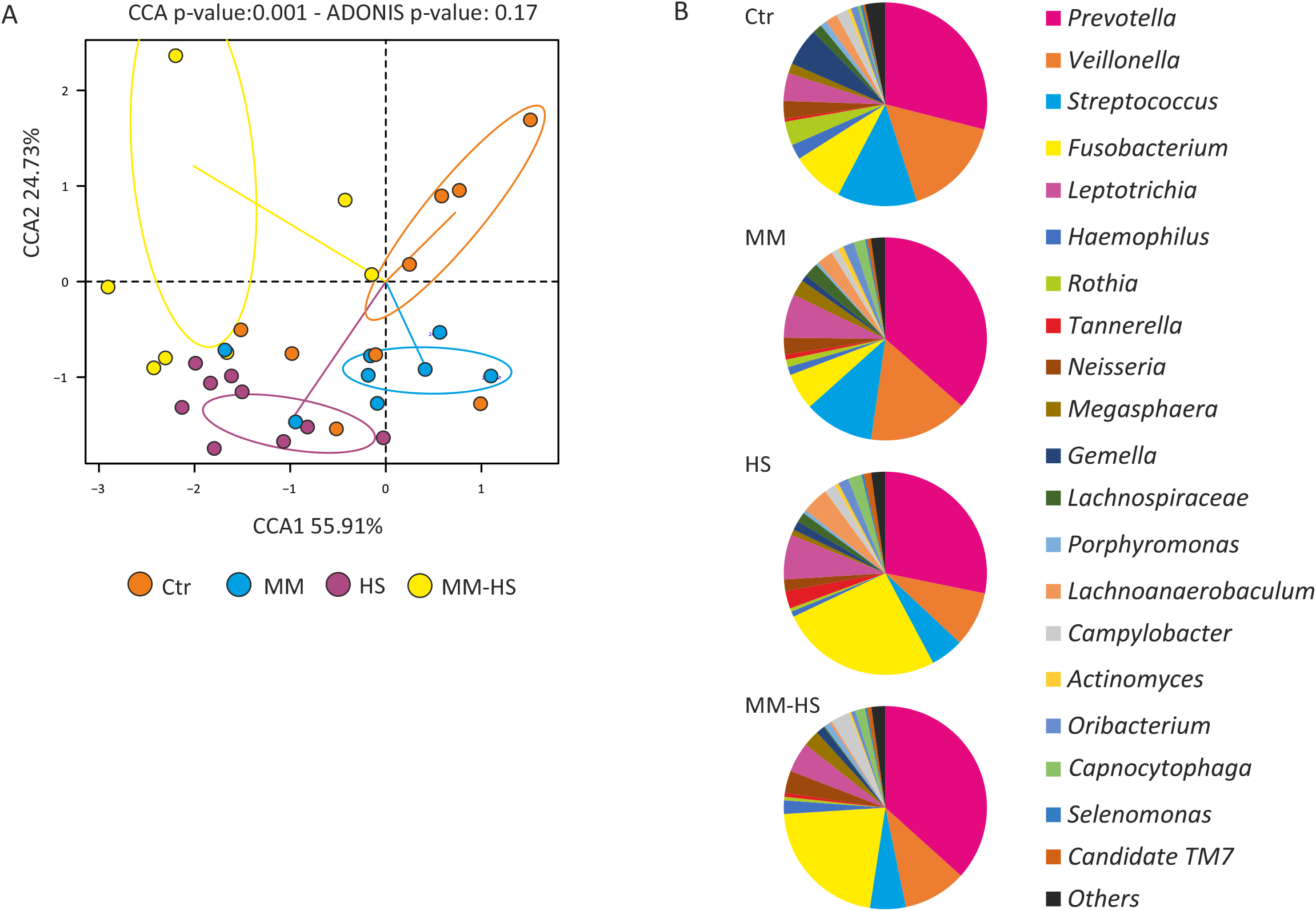
Volatile Sulfur Compounds concentration affects tongue active microbiota composition. A. Bacterial composition at genus level, as determined by assignment of transcripts to oral bacterial genomes, is represented for each group as pie-charts. The median abundance was used for the top 20 most abundant genera. B. Differences between groups were calculated and plotted using Canonical Correspondence Analyses (CCA) and ADONIS test.

The proportion of each taxon was calculated to evaluate the RNA-based composition of the bacterial tongue communities. At the genus level, *Streptococcus* and *Rothia* were more active in *Ctr* while *Fusobacterium* was detected at higher levels in *HS* or *MM*-*HS* groups (**Figure 2B**). At species level, an *unclassified Prevotella* sp., *Streptococcus parasanguinis, Veillonella dispar, Veillonella atypica, Fusobacterium periodonticum, Prevotella histicola, Veillonella sp*., *Streptoccocus salivarius, Rothia mucilaginosa, Prevotella melaninogenica* and an *unclassified Tannerella* sp. together accounted for 50-60% of all bacterial communities within all four groups (see **Suppl. Table 2** for a full list).

When comparing the proportion of RNA-based microbial communities associated to the tongue of halitosis-free individuals and those with elevated VSCs concentrations, several significant differences were found (**Figure 3 and Suppl. Tables 3-5**). As expected, the previously reported H_2_S producer *F. periodonticum* was over-represented in samples with higher concentration of H_2_S (*MM-HS*) in comparison to *Ctr* (**Figure 3**). Even though it was not significant, *F. periodonticum* showed a trend of higher representation (p=0.1) in the *HS* group in comparison to the *Ctr* and *MM* groups. Additionally, other *Fusobacterium* species showed the same pattern as *F. periodonticum*, including *F. nucleatum* and an unclassified *Fusobacterium* sp. (**Suppl. Tables 4-7)**. Interestingly, these species were also significantly over-represented in *MM-HS* in comparison to *MM* and *HS* (**Suppl. Tables 6 and 7**).

**Figure 3.**
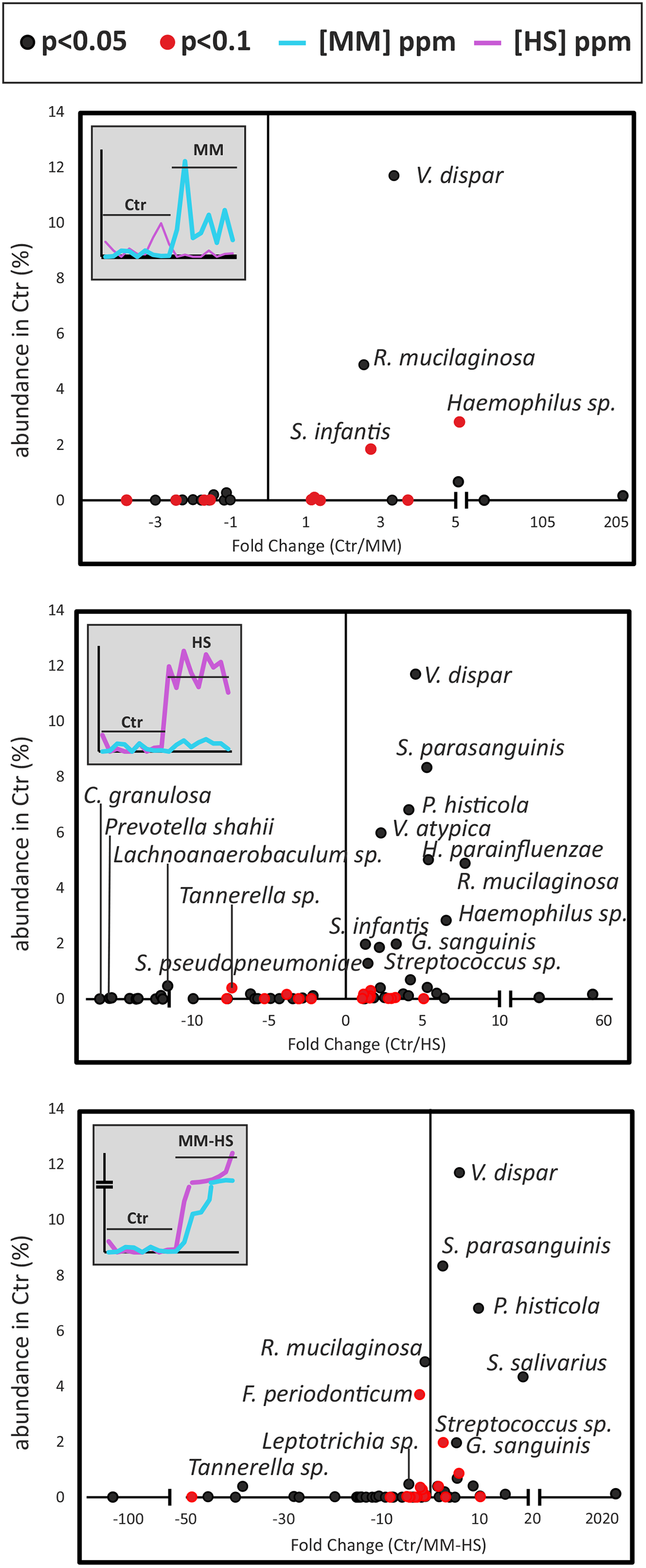
Differences in bacterial composition between control individuals and groups with high concentration of Volatile Sulfur Compounds. Data show the abundance (%) and the fold change (Ctr/corresponding group) for each bacterial species that was significantly different (p<0,05) or showed a tendency (p<0,1) between groups according to DESeq2 analyses. For clarity, only those species with a percentage >1% are indicated. The levels of each VSC for the control (Ctr) and the corresponding halitosis group are shown on the top-left of each panel. A. Ctr vs MM; B. Ctr vs HS; C. Ctr vs MM+HS.

Similarly, several *Prevotella* species were significantly more active in *MM-HS* than in *Ctr*. They included *P. oulorum, P. sacharolytica, P. shahii* and *P. tanneare* (**Suppl. Table 5)**. In addition, other *Prevotella* species were more active in *HS* than *Ctr*: *P. baroniaę P. denticola, P. shahii* and *P. tannerae* (**Suppl. Table 4**). These results suggest an association of active *Prevotella* and *Fusobacterium* species to tongue microbiotas with high concentration of H_2_S. Interestingly, we found *Prevotella histicola* associated with *Ctr* and *MM* patients in comparison to samples with high concentrations of H_2_S (**Figure 3, Suppl. Tables 4-5 and 6-8**). In contrast to *Fusobacterium* and *Prevotella*, several *Streptococcus* species, *Rothia mucilaginosa* and *Veillonella dispar* were under-represented in those patients with higher concentrations of H_2_S (**Figure 3**). In fact, *R. mucilaginosa, Streptococcus paransanguinis* and *V. dispar* were over-represented in halitosis-free individuals in comparison to the three halitosis groups and their mean abundance in this group was remarkably high (4.9, 8.34 and 11.7%, respectively) (**Figure 4**).

**Figure 4.**
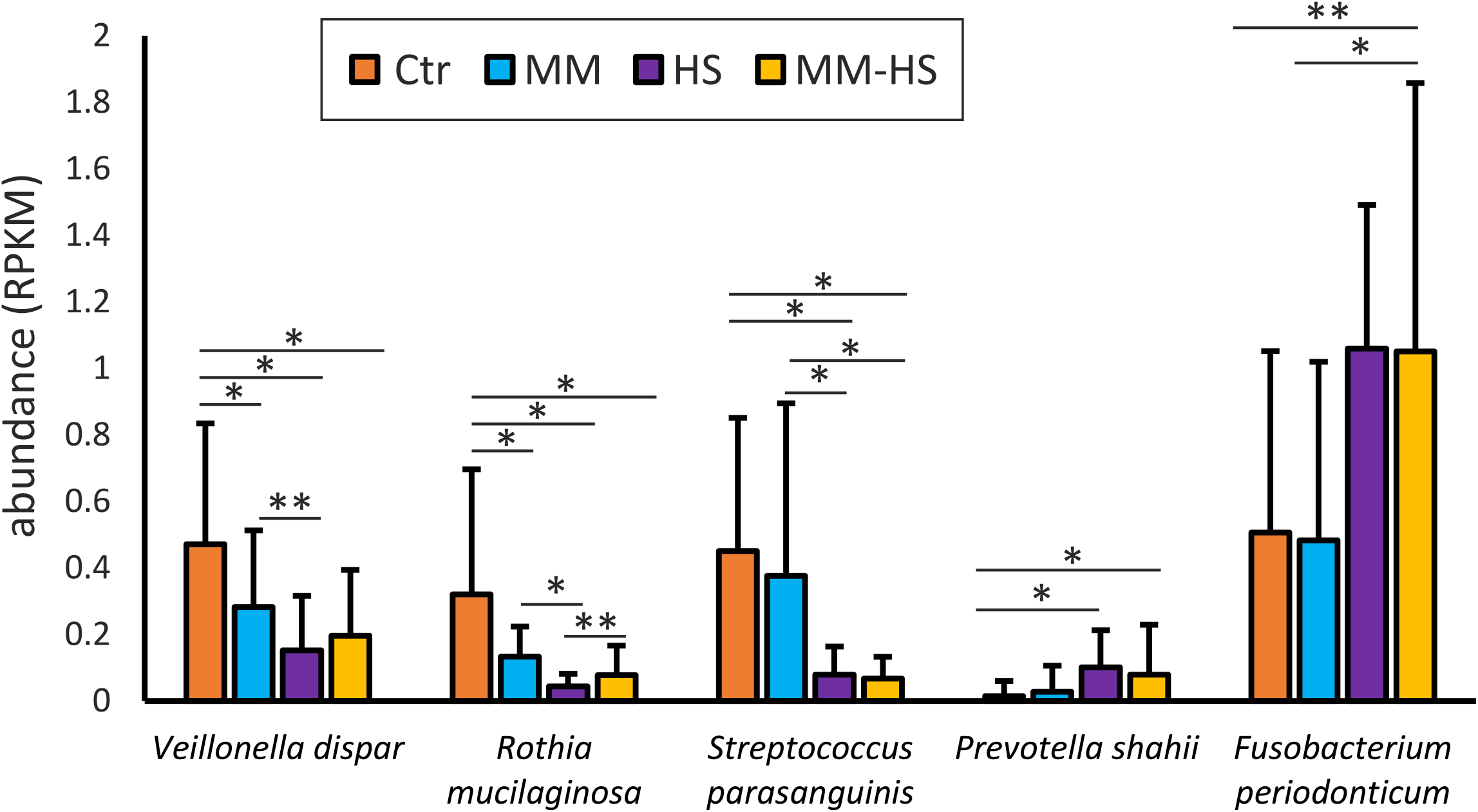
Putative biomarkers of halitosis. Data show the abundance (expressed as number of Reads per gene Length per Million bp, RPKM) of each of the proposed microbial biomarkers for halitosis disease. Statistical differences were highlighted with * (p-value<0,05) or ** (p- value<0,1).

### H_2_S and amino acid biosynthesis in halitosis and halitosis-free patients

Besides the taxonomical composition we also studied the variation in gene expression. CCA plots show that at a functional level the microbiotas had different gene expression profiles depending on H_2_S concentration (**Suppl. Fig. 1**). When comparing different groups with DESeq2, several genes were significantly over- or under-represented (**Sup. Tables 9-14**). In particular, the major differences were found when comparing either *Ctr* or *MM* to either *HS* or *MM-HS* (**Figure 5**). Similar to what we observed using the taxonomical data, the differences between *Ctr* and *MM* (172 differentially represented genes, DRG) were smaller than those between *Ctr* and *HS* (566 DRG) or between *Health* and *MM-HS* (460 DRG).

**Figure 5.**
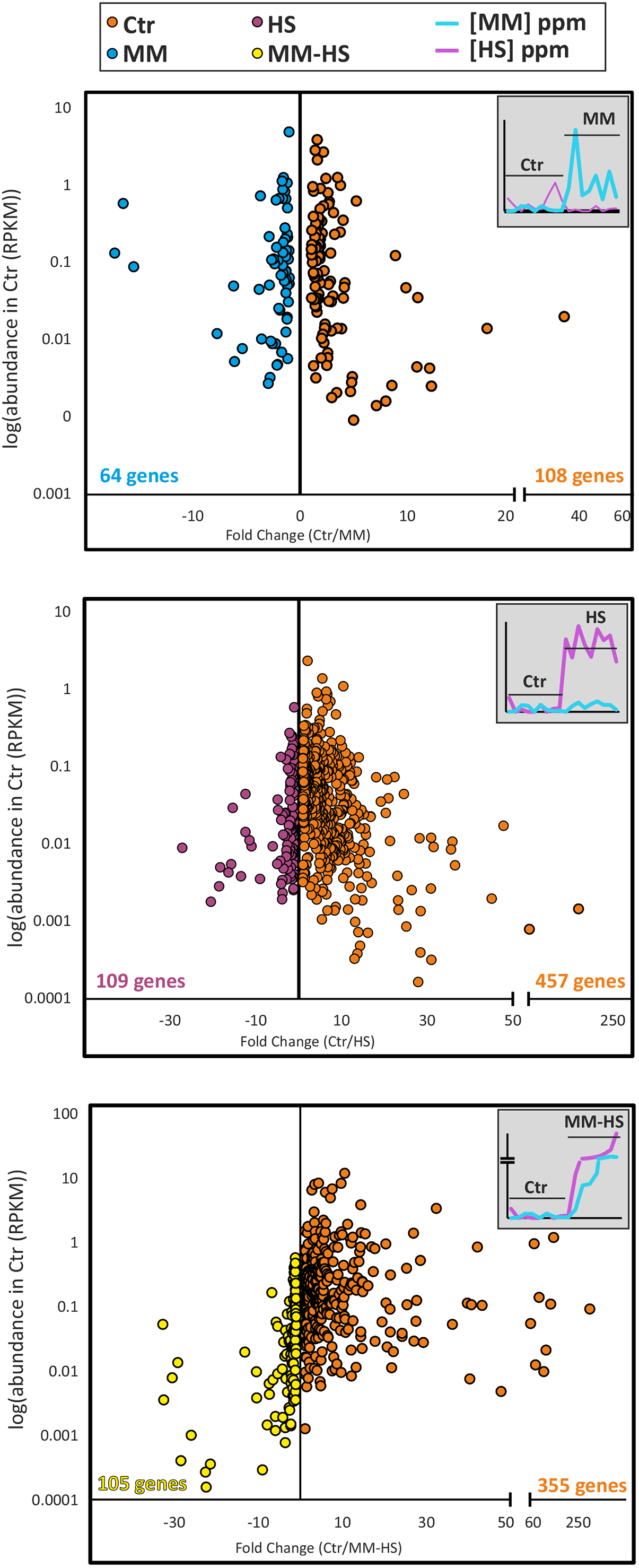
Functional differences between control individuals and groups with high concentration of Volatile Sulfur Compounds (VSCs). The abundance (number of reads normalized by length of the gene and size of the dataset, RPKM) and the fold change (Ctr /corresponding group) for each gene that was significantly different (p<0,05) between groups according to DESeq2 analyses was plotted. The levels of each VSC for Ctr and the corresponding halitosis group are shown on the top-right of each panel. A. Ctr vs MM; B. Ctr vs HS; C. Ctr vs MM+HS.

A major issue relates to whether genes coding for proteins responsible for H_2_S synthesis were over-expressed in *HS* or *MM-HS* groups. According to previous reports, bacteria are able to produce H_2_S using two pathways: reduction of sulfate, and desulphydration of cysteine and methionine [42], each of them presenting two possible routes. In the case of sulfate reduction, there are assimilatory and dissimilatory sulfate reduction pathways (anaerobic) [43]. The desulphydration process includes a reverse trans-sulfuration and a cysteine transamination. Regarding the assimilatory pathway, we only found *cysC* (adenylyl-sulfate kinase) over-represented in the *HS* group compared to *Ctr* out of the 6 genes in this route (**Figure 6A**). Interestingly, the homocysteine desulfhydrase (*mccB*), which is one of the three genes responsible for the L-homocysteine conversion into H_2_S, was over-represented in patients classified as *HS* (**Figure 6A and Suppl. Tables 10 and 12**). There was also a 1.31 fold change difference in *mccB* expression between *MM-HS* and *Ctr* but the difference was not significant (**Figure 6B**).

**Figure 6.**
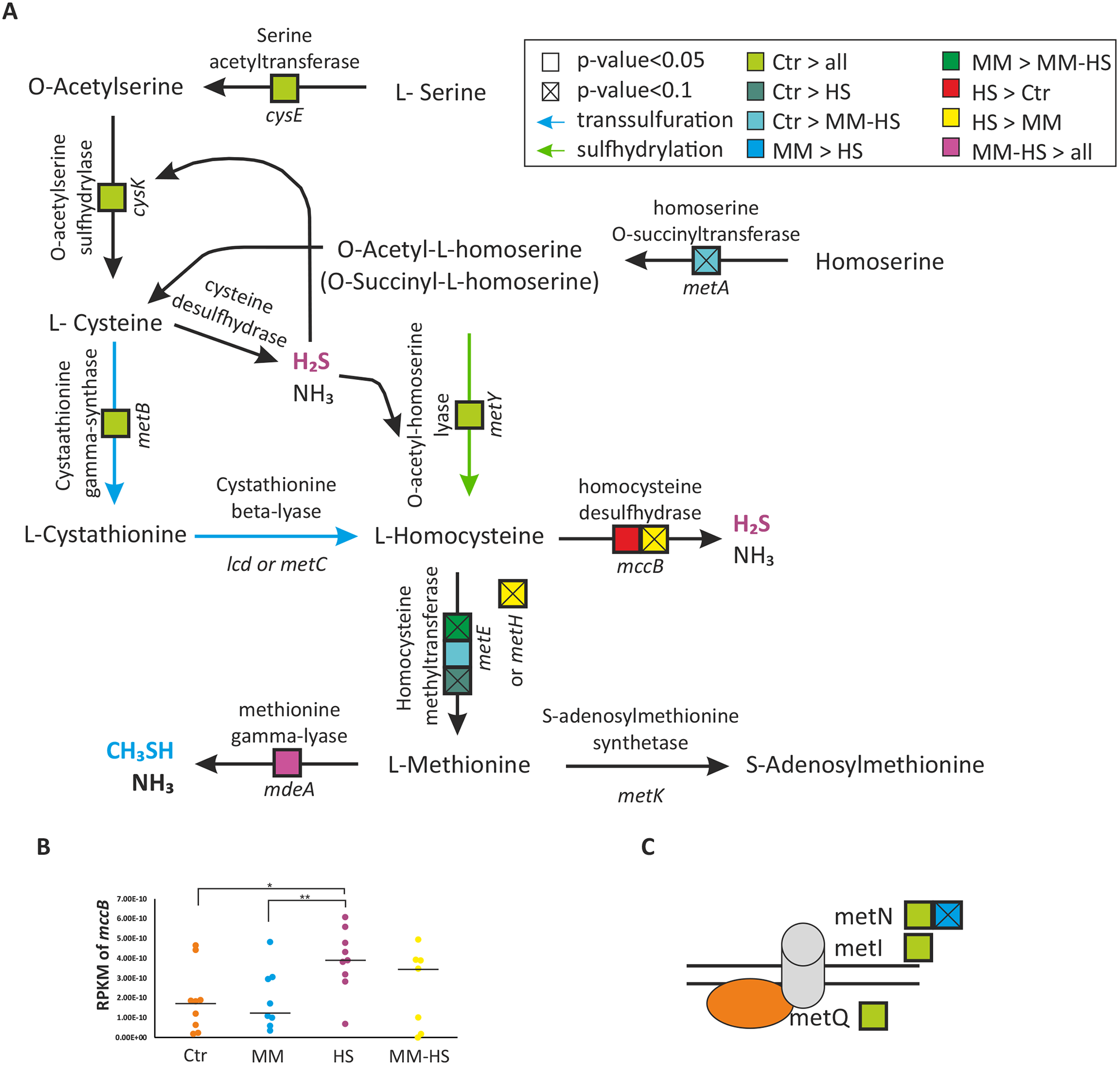
Transcriptional changes of the genes related with the synthesis and transport of different Volatile Sulfur Compounds (VSCs). **A**. The diagram shows the substrate and final products of the enzymatic reactions that lead to the production of hydrogen sulfide and methyl mercaptan. For each gene, a box is shown for each comparison between groups. **B**. An additional diagram is depicted for the *mccB* gene, showing the abundance (RLM) of the gene and the median in each sample. **C**. Transcriptional differences in the VSCs transporters. The proteins involved in the transport of D-methionine to the extracellular medium are plotted. The differences in expression of the genes coding for these proteins are presented in boxes. Same legend was used for A and C.

Even though the concentration of CH_3_SH was high enough to be considered as halitosis in *MM* and *MM-HS* groups, the mean concentration of CH_3_SH in the *MM-HS* group was 2.65 times higher than in the *MM* group (**Figure 1B and Suppl. Table 1**). Among the genes coding for the enzymes responsible for CH_3_SH synthesis, we found *mdeA* (methionine gamma-lyase) over-expressed in *MM-HS* individuals in comparison to the other three groups of samples (**Figure 5A and Suppl. Tables 11**,**13 and 14**). According to this, the increase in CH_3_SH concentration could partly be due to the over-expression of this gene.

Among the genes over-represented in halitosis-free patients, we found the 3 subunits from the D-methionine transport system (*metQIN*) (**Figure 6C and Suppl. Tables 10 and 11**). Interestingly, these 3 subunits were over-represented in halitosis-free individuals in comparison to both *HS* and *MM-HS* groups, suggesting an important role in oral health. Interestingly, *metY* (O-acetylhomoserine [thiol]-liase), which is responsible for the formation of L-homocysteine using H_2_S, was over-represented in *Ctr* samples when compared to *HS* and *MM-H* (**Figure 6A and Suppl Tables 10 and 11**). Moreover, the utilization of sulfide as a sulfur source to form L-cystathionine was also favored in *Ctr* as indicated by the over-representation of *cysE* (Serine acetyltransferase), *cysK* (cysteine synthase) and *metB* (cystathionine gamma-synthase) in comparison to *HS* and *MM-HS* groups.

The metatranscriptomic data would therefore indicate that the formation of sulfide would be more intense in HS and MM-HS patients whereas halitosis-free individuals would potentiate the utilization of sulfide for aminoacid production.

### Nitrate reduction is under-expressed in Halitosis patients

Genes involved in the reduction of nitrate (*narHGIJW*) into nitrite were over-expressed in *Ctr* and *MM* groups in comparison to the *HS* group (**Figure 7)**. Apart from these, the genes coding for nitrogen (N_2_) fixation into ammonium (*nifX*) and the reduction of nitrite into ammonium (*nirB* and *nirD*) were also over-expressed in halitosis-free individuals in comparison to *MM-HS*. Another gene for nitrite reduction into ammonium, *nrfA*, was detected, but did not differ significantly between the groups. In summary, reduction of nitrate into nitrite and the formation of ammonium appears to be favored in the oral cavity of halitosis-free individuals. This agrees with the taxonomic analysis described above where two nitrate reducing species, *Rothia mucilaginosa* and *Veillonella dispar*, and a third species that has also shown to produce nitrite (possibly by nitrate reduction), *Streptococcus parasanguinis* [44], were more active in halitosis-free individuals.

**Figure 7.**
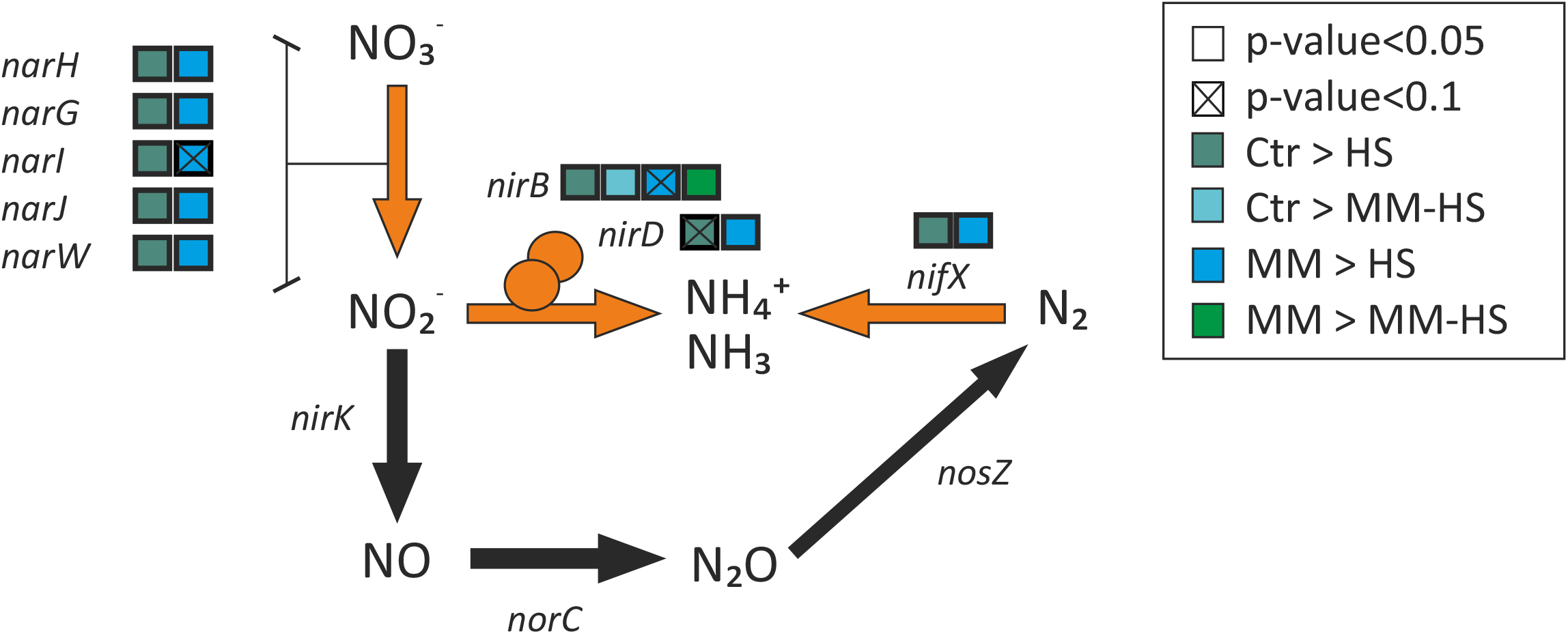
Transcriptional changes of the genes related with nitrate metabolism. The diagram shows the substrates and final products of the enzymatic reactions that lead to denitrification and the production of ammonium from nitrite and nitrogen. For each gene, a box is shown for each comparison between groups and orange arrows highlight reactions with significant variations between groups.

### Classifying halitosis patients based on the VSC profile

This is the first study including a comparison in tongue microbial composition and activity between halitosis-free subjects with patients with high concentrations of CH_3_SH only (MM group) but not H_2_S, or with patients with high concentrations of H_2_S only (HS group). Surprisingly, 5 out of 9 *Ctr* samples clustered within the *MM* group in the CCA at the species level (**Figure 2A**). At a functional level, the differences between *Ctr* and *MM* groups were even lower (**Suppl. Fig. 1**). Moreover, the number of species (**Figure 3**) and genes (**Figure 5**) differentially represented between patients with high concentration of only *CH*_*3*_*SH* and *Ctr* was noticeably low in comparison to *Ctr* vs *HS* or *Ctr* vs *MM-HS*. Finally, CCA plots showed the *HS* group significantly separated from *Ctr* and *MM*, especially at the gene level, suggesting that H_2_S but not CH_3_SH could produce a significant change in the active microbiota.

## Discussion

In the present study, we have analyzed the composition and gene expression of the active tongue microbiota in halitosis-free individuals and halitosis patients using a metatranscriptomic approach. As this condition is the consequence of VSCs production, we measured the concentration of these gases and placed volunteers in four groups based on their VSC profiles. Independently of the group, the composition of the tongue microbiota detected in this study presented similarities to previously reported tongue communities by other authors [24,45,46]. Interestingly, a recent spatial-ecological study of the tongue-dorsum microbiota found a lower proportion of *Prevotella* and *Fusobacterium* than our work [47]. However, this study analyzed DNA instead of RNA, indicating that there these two bacterial genera could be proportionally more active than previously anticipated.

The comparison of active bacteria between halitosis-free individuals and those with high concentration of CH_3_SH and/or H_2_S supported previous studies in which known VSCs producers (e.g. *Porphyromonas, Fusobacterium* and *Prevotella* species) were associated with a high concentration of H_2_S [5,48–50]. Moreover, we detected an association of halitosis-free individuals with common oral commensals such as representatives of *Streptococcus, Corynebacterium, Rothia* and *Veillonella*. In agreement with this, Seerangaiyan et al. (2017) found an increase in several *Streptococcus* spp. and *Rothia dentocariosa* in halitosis-free individuals compared to individuals with halitosis [51].

One of the main findings in our analysis was the correlation of bacterial activity profiles with different VSC levels. According to the differences observed in active microbial proportions, the increase in H_2_S concentration was related to clear taxonomic changes associated to disease (i.e., dysbiosis), while this bacterial composition shift was modest when only CH_3_SH was elevated. In fact, patients with an increase in CH_3_SH but not H_2_S did not differ significantly in microbiota composition compared to halitosis-free individuals. We hypothesize that CH_3_SH in halitosis is a secondary metabolite of tongue-associated microbiota, but the presence of CH_3_SH by itself may not be an indication of dysbiosis.

Interesting similarities were found between our results and those of Takeshita and colleagues (2012), who also classified patients in different groups based on their VSCs profiles, but used bacterial characterization by 16S rRNA pyrosequencing in saliva samples [15]. For example, *Porphyromonas and Fusobacterium* were associated in both studies with higher concentration of H_2_S. Similarly, *Veillonella* was found associated with the group with higher concentration of CH_3_SH in comparison to those with H_2_S. A difference was found when comparing the *MM* group with *Ctr* in our study, namely that *Veillonella* species were more active in halitosis-free individuals, but in their study more *Veillonella* was found in the MM group. Additionally, Takeshita *et al*. associated *Neisseria* with higher concentration of H_2_S. In our study, some species of *Neisseria* (with highest similarity to *N. lactamica, N. mucosa, N. gonorrhoeae* and *N. meningitidis*) were more active in *HS* and *MM-HS* groups whereas others were more active in *MM* individuals compared to *HS* and *MM-HS* (*N. subflava, N. bacilliformis* and *N. polysaccharea*).

Previous epidemiological reports have observed a correlation of halitosis with periodontitis [2,18,52]. Both patients with halitosis and patients with gum disease have more tongue coating than halitosis-free individuals [5,52]. In our dataset, we did not find differences in tongue coating between halitosis-free and any of the 3 halitosis groups (p>0.1). Remarkably, there are several periodontal pathogens among the mentioned species associated with malodor found in this study, such as representatives of *Porphyromonas, Fusobacterium* or *Prevotella*. These results in active microbial communities support the idea of an overlap in the bacterial composition of halitosis and periodontitis, and a possible link between these diseases [18,52].

The periodontitis-associated genera that were higher in the halitosis groups (*Porphyromonas, Fusobacterium* and *Prevotella*) included species which are known to produce VSCs (i.e. *Porph. gingivalis, F. nucleatum*, and *Prev. intermedia*) [27,53] and have been commonly detected in higher amounts in the tongue of halitosis patients [5,24,51]. Thus, we propose these species as potential universal halitosis biomarkers. On the other hand, we detected *Veillonella dispar, Streptococcus parasanguinis* and *Rothia mucilaginosa* significantly more active in individuals classified as halitosis-free. These are commensal members of the oral microbiota and *Streptococcus* and *Rothia* have previously been detected in higher proportions in groups with lower VSCs concentration and organoleptic score [54]. Interestingly, *Veillonella* species have been found in dental cavities and have been reported to produce H_2_S *in vitro* [55]. In our study, *dispar* was more active in individuals that did not produce significant amounts of H_2_S. A possible explanation is that in halitosis-free *in vivo* conditions, *V. dispar* does not produce H_2_S, while in disease conditions, other species (e.g., *Fusobacterium* spp. and *Prevotella* spp.) use cysteine and/or reduce sulfate more efficiently. It could also be possible that *V. dispar* is producing VSCs but at a concentration below the established thresholds or that in the presence of nitrate, sulfate reduction is inhibited by more energy-efficient nitrate reduction [56]. The higher activity of *R. mucilaginosa*, which is a highly efficient nitrate reducer [57], in halitosis-free individuals, also suggests that nitrate could favor health-associated conditions. Taken together, our data suggest that nitrate could have a beneficial effect on tongue communities involved in halitosis, as previously suggested [58]. This agrees with the well-known phenomenon that reducing nitrate provides more energy than reducing sulfate, and therefore microorganisms normally switch to denitrification when nitrate is available, which could therefore imply lower VSC production [56]. Secondly, nitrite is reduced to nitric oxide by several oral bacteria, and nitrite reductases have been shown to use hydrogen sulfide as electron donor, which would therefore lower the levels of H_2_S in the oral environment [59]. Another indirect suggestion comes from sewage treatment plants, where nitrate can be used as a biological agent to treat malodor by limiting microbial VSCs production resulting from sulfate reduction [56]. Therefore, we propose *V. dispar, S. parasanguinis* and *R. mucilaginosa* as biomarkers for halitosis-free individuals and suggest nitrate as a potential prebiotic to promote a halitosis-free microbiome. Future experimental and clinical work should confirm this hypothesis.

## Conclusions

This is the first report of microbial gene expression profiles in the tongue biofilm of halitosis patients. The comparison of these with halitosis-free individuals indicated that an elevated concentration of H_2_S, or its combination with CH_3_SH, induced dysbiosis in the tongue microbiota. The changes in active microbiota composition associated with an increase in VSCs concentration are in accordance with previous reports. These include an increase in *Prevotella, Fusobacterium* and *Porphyromonas* and a reduction in commensal oral members such as *Streptococcus, Veillonella* and *Rothia*. Our metatranscriptomic data provide the first evidence that genes involved in VSCs production are being expressed *in vivo* and identify several routes by which the bacterial communities in halitosis-free individuals consume VSCs to transform them in non-halitosis compounds. Finally, we have detected a relationship between halitosis and a repression of genes responsible for nitrate reduction. Thus, we propose those pathways involved in VSC consumption and in nitrate reduction as potential therapeutic targets for preventing and treating the disease.

## Data Availability

All data produced in the present study are available upon reasonable request to the authors. The datasets generated during the current study are available in the SRA repository with the accession number PRJNA735705.

## Acknowledgements

Authors would like to thank the FISABIO Sequencing Platform and particularly Nuria Jimenez for their collaboration.

## Authors’ contributions

AM and BR design the study. SLl and CLl obtained the tongue samples and measured the VSCs concentration using the OralChroma. MCD performed the RNA extractions, analyzed the sequencing data and performed the statistical analyses. MCD, BR and AM interpreted the data and wrote the manuscript.

## Consent for publication

Not applicable

## Availability of data and materials

The datasets generated during the current study are available in the SRA repository with the accession number PRJNA735705.

## Figure captions

**Supplementary figure 1. Canonical Correspondence Analyses plot of gene function profiles, performed at genes level according to the KEGG database classification**. Differences between groups were calculated and plotted using Canonical Correspondence Analysis (CCA) and ADONIS test.

