## Supplementary figures and images for "The tongue biofilm metatranscriptome identifies metabolic pathways associated with halitosis and its prevention"

### Suppl. Fig. 1

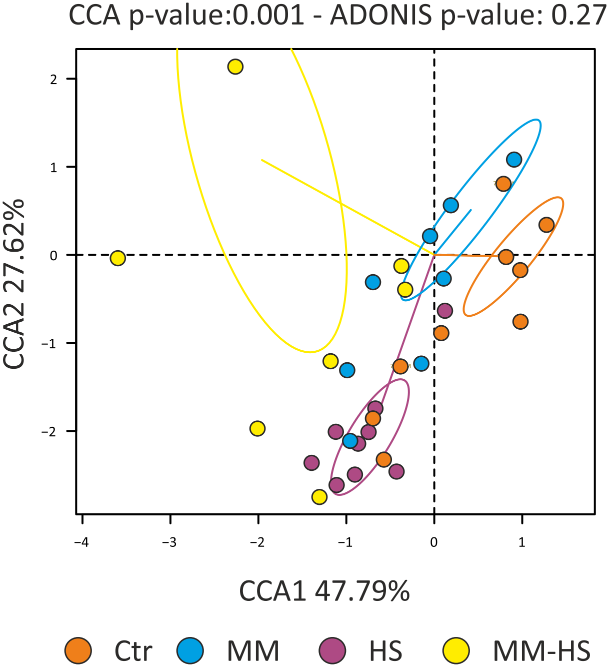
